# Tacrolimus variability and creatinine predict readmission after liver transplantation

**DOI:** 10.64898/2026.07.02.26357106

**Authors:** Kevin Korenblat

## Abstract

Unplanned readmissions after liver transplantation occur in over 30% of recipients, yet no validated prediction models exist, and prior observational studies suffer from immortal time bias. The optimal readmission window for outcome prediction and the feasibility of early risk stratification remain undefined. This study is a retrospective analysis of 922 adult liver transplant recipients (August 2018-August 2025) at a single center. Time-varying Cox regression evaluated 14-, 30-, and 90-day readmission windows as predictors of 1-year mortality, correcting for immortal time bias. Gradient-boosted machine learning models leveraging 528,400 laboratory measurements (28 analytes) predicted 90-day readmission using either complete hospitalization data or data restricted to postoperative day 7. Feature importance was quantified by gain, and clinical utility was assessed through risk stratification. Among 902 hospital survivors, 342 (37.9%) experienced an unplanned readmission within 90 days of initial discharge. Only the 90-day readmission window predicted 1-year mortality in time-varying analysis (HR 1.73, 95% CI 1.17-2.57, p=0.006). The model for readmission using complete data achieved AUC 0.614 (95% CI 0.576-0.652); the postoperative day 7 restricted model achieved AUC 0.615 (95% CI 0.577-0.652), with no meaningful performance difference. The tacrolimus coefficient of variation × peak creatinine interaction was the dominant predictor in both the complete model (17.3% importance, rank 1) and the day 7 restricted model (20.4% importance, rank 2). This interaction stratified patients into high-risk (tacrolimus CV >0.3 and creatinine >2.0 mg/dL; 49.8% readmission) versus low-risk (24.8% readmission) groups (risk ratio 2.01, p<0.001). These results identify a modifiable biological determinant of readmission and establish a framework for targeted interventions to reduce unplanned readmission and improve post-transplant outcomes.

## INTRODUCTION

Unplanned readmissions after liver transplantation are an outcome metric of value to patients, providers, and payers[1]. Reported rates of readmission are at least 30% and the risk is associated with education level, age, discharge location, and renal function[2–6].

Research into post-liver transplant readmission, however, has significant limitations which include no agreed upon time interval within which a readmission is associated with a relevant clinical outcome. Though most studies use thirty- or ninety-days, those choices are not evidence-based. The research into readmissions also frequently fails to account for an immortal time bias[7,8], and there are no studies that have attempted a predictive model for readmissions.

The absence of prediction models stems from both methodological and practical challenges related to the fact that relevant features may not be captured in a way that allows for efficient abstraction. A way to navigate this issue is to apply modern analytic techniques to the most accessible data. Current electronic health records (EHR) systems capture thousands of timestamped laboratory measurements and store this data in relational databases[9]. The amount of data is vast, yet health care providers typically use only a fraction of this information and, often, only at static time points for clinical decision making, including prognostic modeling. These limitations can be overcome using contemporary machine learning methods which can harness high-dimensional data to identify patterns invisible to traditional regression-based approaches[10].

The purpose of the study was twofold. First, the study sought to clarify the time interval for unplanned readmissions most relevant to post-transplant outcomes using methods free of immortal time bias. Second, the study leveraged 528,400 laboratory measurements across 902 patients (approximately 573 measurements per patient across 28 analytes) to develop machine learning models to predict unplanned, post-liver transplant readmissions using only data available by post-operative day (POD) 7.

## METHODS

### Study Population

The study is a single-center, retrospective analysis of all adult (≥18 years) liver transplant recipients, including simultaneous kidney-liver, heart-liver, and lung-liver transplants, performed between August 4, 2018, and August 13, 2025, at Barnes-Jewish Hospital (BJH). Patients undergoing re-transplantation were excluded. The study start reflects the implementation of the EPIC EHR (Epic Systems Corporation, Verona, WI) at BJH. The Washington University School of Medicine Human Research Protection Office approved the study with waiver of informed consent.

### Data Collection

All laboratory measurements from hospital admission to discharge during the index hospitalization that transplantation was performed were extracted from the EHR. Measurements spanned 28 unique analytes include complete blood count, comprehensive metabolic panel, coagulation studies, arterial blood gases, liver function tests, and immunosuppression drug levels at trough (tacrolimus and cyclosporine). Timestamp was recorded in hours relative to operating room entry. Data on the readmission diagnosis was also extracted from the EHR.

Patient demographics, outcomes, and donor age were extracted from the Organ Procurement Transplant Network Standard Transplant Analysis and Research file and included as static predictors in the models. Multi-organ transplant status was captured as a binary indicator and as an interaction term with creatinine and bilirubin trajectories to capture the distinct recovery dynamics of simultaneous liver-kidney recipients.

### Outcomes

The primary outcome was unplanned readmission within 90 days of first discharge among 902 survivors who did not undergo retransplantation during the index admission.

### Feature engineering

Laboratory data were aggregated into clinically defined time windows: baseline (pre-transplant), intraoperative (0-6 hours), post-operative (6-24 hours), POD 1 (24-48 hours), and POD 0-7 (0-168 hours). For each analyte and window, the mean, maximum, most recent, and timeframe of reaching maximal value were utilized.

Derived features included: Model for End Stage Liver Disease (MELD) scores at baseline, POD 0, and POD 7; trajectory features capturing lab trends, and interaction terms include aspartate aminotransferase (AST) x recipient age, donor age x MELD, and multi-organ status x organ function trajectories. The tacrolimus coefficient of variation (CV) was calculated as the ratio of standard deviation to mean trough level drawn during POD 0-7 (minimum of two measurements required).

For the POD 7 model, all features were restricted to data available by POD 7 (168 hours). Length of stay, discharge disposition, and any laboratory data from after POD 7 were excluded.

### Model Development

Gradient-boosted decision trees using XGBoost[11] (version 3.2.0.1) were implemented. Missing values were imputed using within-training-set median imputation. Features with >50% missingness or variance <0.01 were removed. Regularization was applied to prevent overfitting given low event rates: max_depth=3, eta=0.01, min_child_weight=20, gamma=1, lambda=2, alpha=1. Scale_pos_weight was set to the inverse event rate to handle class imbalance.

### Outcome definition validation

To validate the 90-day readmission outcome and compare readmission windows, the study included Cox proportional hazards regression with time-varying covariates, examining readmissions at 14-, 30-, and 90-days post-discharge as predictors of 1-year all cause graft loss. The time varying covariate approach was used to correct for immortal time bias. Each patient contributed person-time to the “not yet readmitted” stratum from discharge until readmission, then switched to the “readmitted” stratum for the remainder of the follow-up to eliminate the survival requirement artifact inherent in native binary readmission models. All Cox models were adjusted for recipient age, gender, donor age, and multi-organ status. The uncorrected analysis was also performed for comparison. Additionally, the Wilson score method was used to estimate the proportion of readmissions that were truly unplanned in the full cohort based on manual chart review validation.

### Statistical analysis

Model discrimination was evaluated using 5-fold cross-validated areas under the curve (AUC) with stratified sampling. Mean AUC ± standard deviation with 95% confidence intervals calculated using the t-distribution is reported. Feature importance was quantified by gain. The prospective performance loss (ΔAUC) was calculated as the proportional decrease from the model incorporating all hospitalization data. Analysis were conducted in R version 4.5.4[12].

Model calibration was assessed using Hosmer-Lemeshow goodness-of-fit testing (10 groups) and visualized with calibration curves. The Brier score was calculated as the mean squared error between predicted probabilities and observed outcomes.

### Software and Analytical Support

R code for feature engineering, XGBoost model implementation, and data visualization was developed with assistance from Claude Sonnet 4.6 (Anthropic, San Francisco, CA), a large language model used as a coding assistant. All generated code was reviewed line-by-line, tested against the dataset, and modified as needed by the author. Data extraction, cleaning, analysis, interpretation of results, and all scientific conclusions were performed independently by the author without AI assistance.

## RESULTS

### Transplant cohort

The 922-patient transplant cohort (Table 1) included 816 liver transplants (88.5%) and 106 multi-organ recipients (11.5%): 103 simultaneous liver-kidney (11.2%), 1 heart-liver, and 2 lung-liver. Median recipient age was 60 years (IQR 52-67), and 62% were male. Median donor age was 50 years (IQR 35-61). The median length of stay was 8 days (IQR 6-13) and 70% of the subjects remained hospitalized by POD 7 (Supplementary Figure 1). Donors included 752 (81.6%) donations after brain death, 166 (18%) donations after circulatory death, and 4 (0.4%) living/domino donors.

**Table 1.**
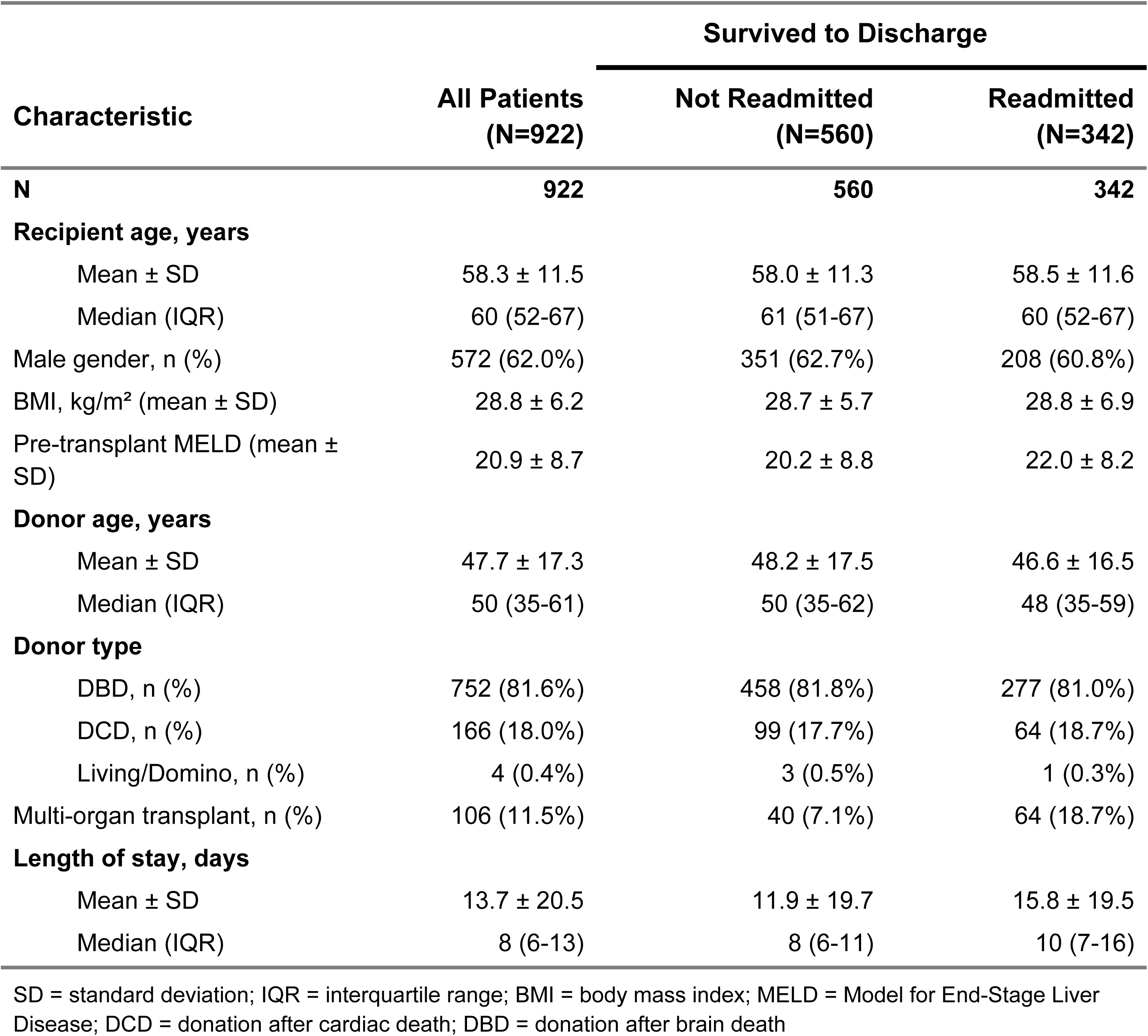
Baseline Recipient and Donor Characteristics, Stratified by 90-Day Readmission Status.

902 patients survived the index admission to discharge and 342 experienced unplanned readmissions within 90 days of discharge. To validate the electronic health record classification of unplanned readmissions, charts of 50 randomly sampled patients (25 readmitted within 30 days, 25 from 31-90 days) were manually reviewed. All 50 (100%) were confirmed to be unplanned readmissions. The Wilson score method provides 95% confidence that at least 92.9% of readmissions in the full cohort (≥318 of 342) were unplanned.

Outcome Validation: Readmission Window and 1-Year Mortality A time-varying covariate Cox regression was performed to determine the appropriate readmission window and validate the 90-day outcome. Readmission intervals of 14, 30, and 90 days were explored as predictors of 1-year patient mortality, adjusting for recipient age, gender, donor age, and multi-organ transplant status. This approach corrects for immortal time bias, whereby patients must survive to discharge and long enough post-discharge to qualify for readmission. Absent this correction, the analysis indicated readmission was protective for survival (14-day HR 0.746, 95% CI 0.424-1.31, p=0.311).

After correction, a pattern to the association of readmission with one year patient mortality emerged (Figure 1). Neither 14-day (HR 0.781, 95% CI 0.443-1.38, p=0.392) nor 30-day readmission (HR 1.03, 95% CI 0.653-1.62, p=0.908) were associated with one year mortality. In contrast, the 90-day readmission was a strong and highly significant predictor of 1-year mortality (HR 1.73, 95% CI 1.17-2.57, p=0.006).

**Figure 1.**
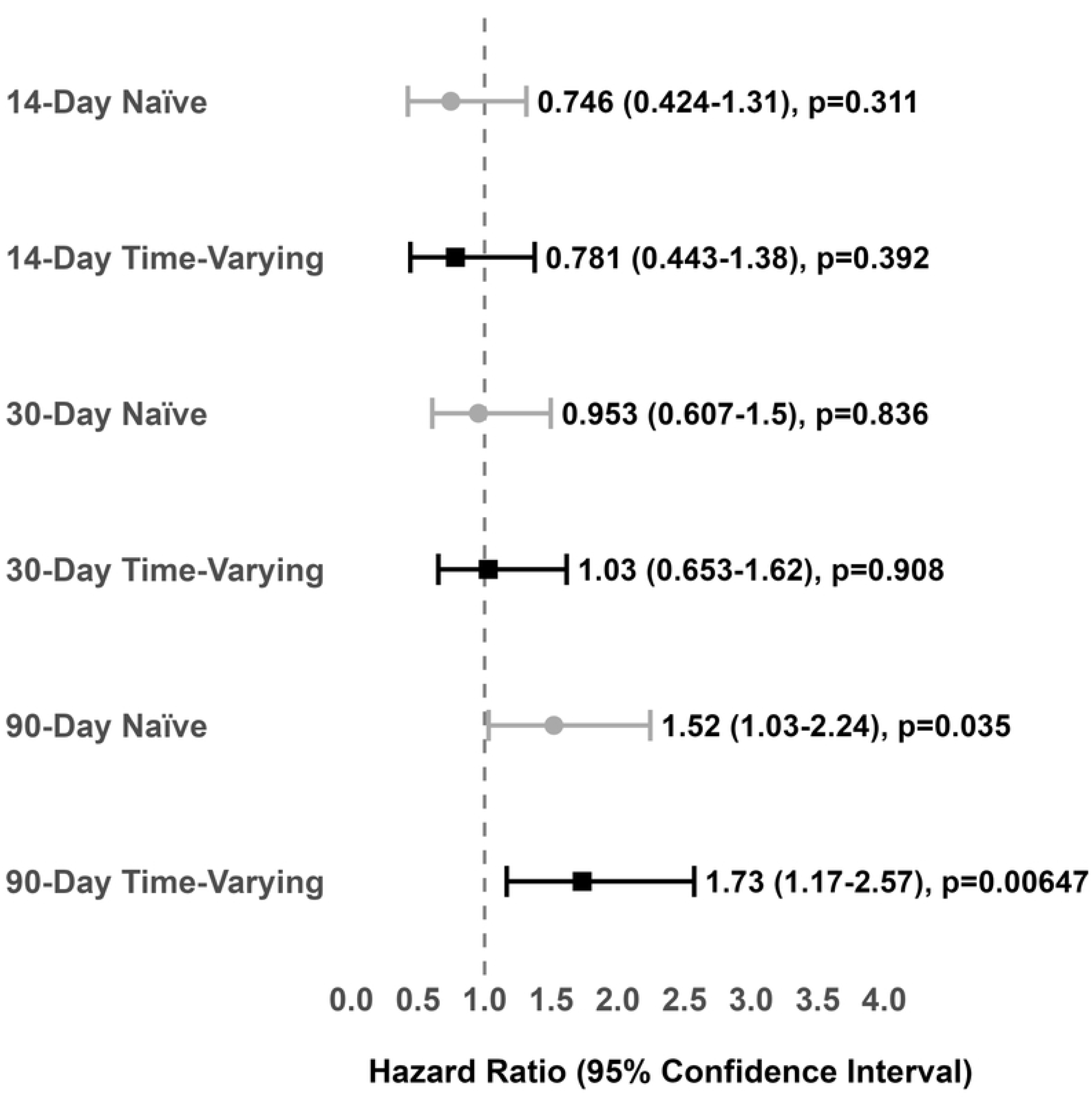
Forest plot comparing naïve and time-varying Cox proportional hazards models. Models are for the association between readmission (at 14-, 30-, and 90-day windows) and 1-year patient mortality. Models adjusted for recipient age, gender, donor age, and multi-organ status.

### Model Performance

Two models of readmission were created. The model incorporating all available inpatient data, including LOS and discharge disposition, achieved an AUC 0.614 (95% CI 0.576-0.652) with stable performance across cross-validation folds (mean CV AUC 0.619 ± 0.065). The POD 7 restricted model achieved an overall AUC 0.615 (95% CI 0.577-0.652) with stable performance across cross-validation folds (mean CV AUC 0.623 ± 0.038) with no meaningful performance difference (Figure 2).

**Figure 2.**
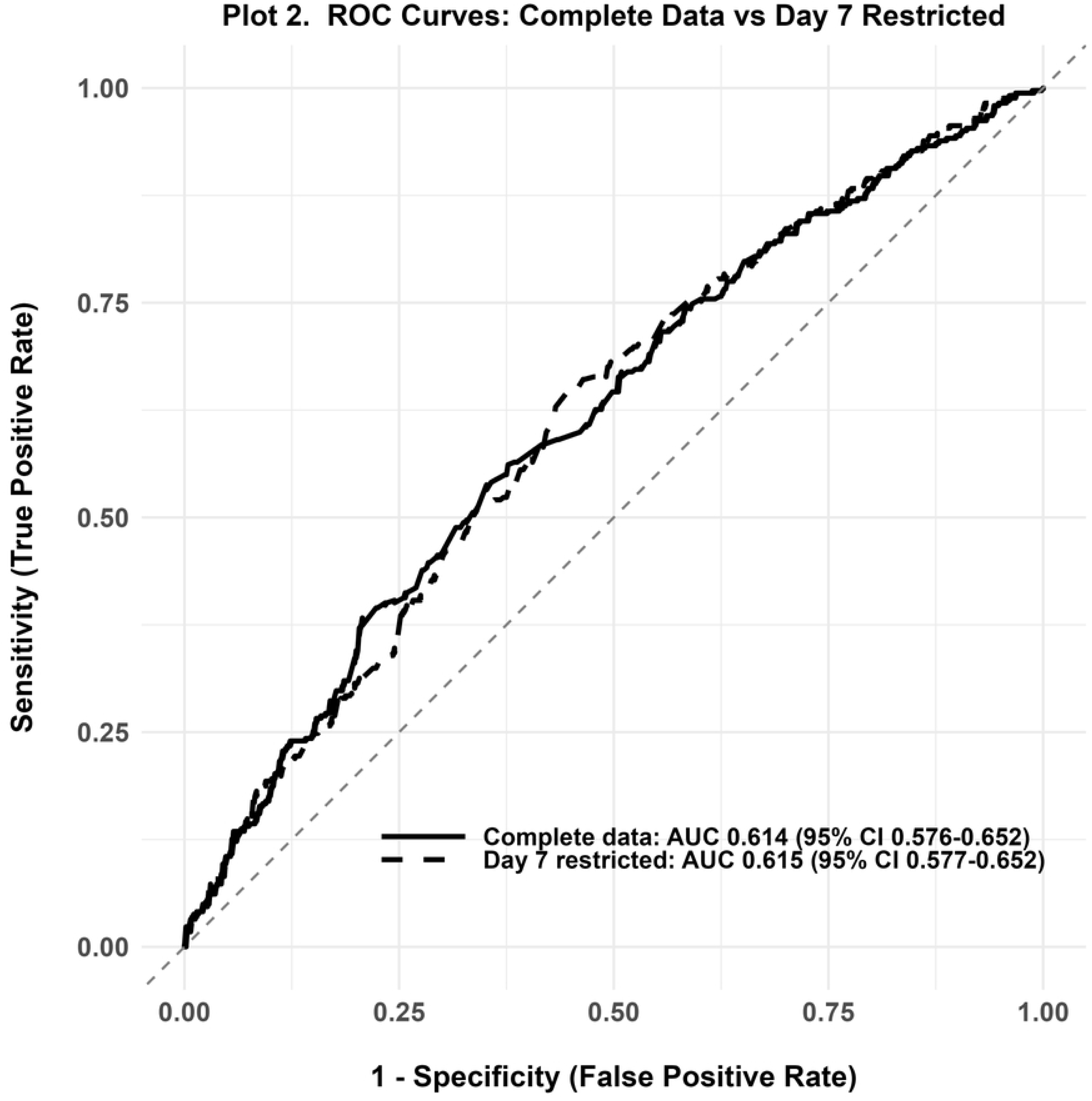
ROC curves for predictive models. Models for readmission using either complete data (solid line) or day 7 restricted data (dashed line).

Calibration analysis demonstrated that predicted probabilities aligned well with observed readmission rates. Calibration plots showed strong agreement between predicted and observed readmission rates across the middle range of predicted probabilities (20-60%), with minimal deviation from the diagonal reference line.

Calibration was less precise in the lowest and highest deciles, reflecting smaller sample sizes and lower event counts in these extreme risk categories (Figure 3). Brier scores were 0.240 and 0.241 for the complete and POD 7 restricted models, respectively, indicating good overall probabilistic accuracy. (Supplementary Table 1).

**Figure 3.**
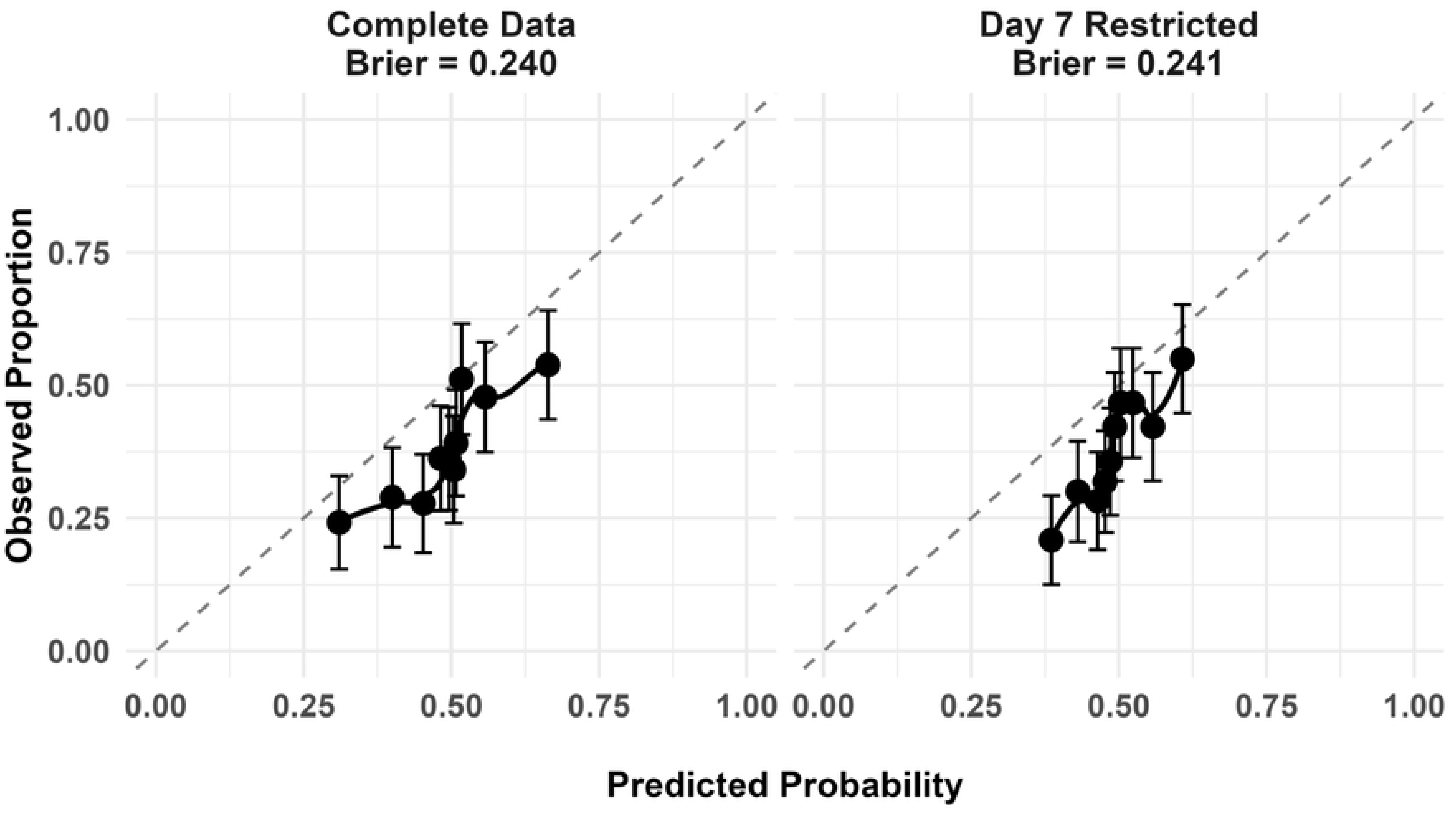
Calibration plots. Plots show predicted versus observed readmission rates across deciles of predicted risk for complete model (left) and day 7 model (right). Points represent decile bins, dashed line represent perfect calibration.

At the standard threshold of 0.5, the POD 7 model achieved sensitivity of 46.2%, specificity of 68.9%, positive predictive value 47.6%, and negative predictive value 67.7%. (Supplementary Table 1).

Feature importance analysis revealed substantial overlap between the complete and POD 7 restricted models (Table 2). Recipient BMI accounted for 14.5% (rank 2) and 21.3% (rank 1) in the complete model and day 7 restricted model, respectively.

**Table 2.** Model Features and Importance.

| Rank | Complete Model |  | Day 7 Restricted Model |  |
| --- | --- | --- | --- | --- |
|  | Feature | Importance | Feature | Importance |
| 1 | **Tacrolimus CV × Creatinine** | **17.3%** | Recipient BMI | 21.3% |
| 2 | Recipient BMI | 14.5% | **Tacrolimus CV × Creatinine** | **20.4%** |
| 3 | AST Slope | 7.6% | Creatinine Variability (SD) | 11.4% |
| 4 | Bilirubin Variability (SD) | 6.8% | Creatinine Peak Timing | 6.5% |
| 5 | AST × Recipient Age | 6.1% | Creatinine Rise Rate | 5.3% |
| 6 | Creatinine Peak Timing | 5.5% | Bilirubin Variability (SD) | 4.6% |
| 7 | Multi-organ Transplant | 5.5% | Mean Tacrolimus Level | 4.6% |
| 8 | Total Bilirubin (POD 0) | 5.4% | Donor Age | 3.7% |
| 9 | Number of tacrolimus measurements | 5.0% | AST × Recipient Age | 2.2% |
| 10 | Creatinine Variability (SD) | 4.3% | Multi-organ Transplant | 2.2% |

Features ranked by gain metric from gradient-boosted decision trees

Features related to creatinine levels and tacrolimus level variability accounted for 27.1% and 43.6% of model importance, respectively.

The interaction between tacrolimus CV and peak creatinine emerged as the dominant predictor in both models: 17.3% in the complete model (rank 1) versus 20.4% in the POD 7 restricted model (rank 2). Hepatic injury dynamics were important in both models. In the complete model, AST slope accounted for 7.6% of importance (rank 3) to the complete data model and both models had AST x recipient age as a feature (complete model 6.1%; day 7 restricted model 2.2%).

To evaluate the clinical relevance of the tacrolimus CV x peak creatinine interaction, patients were stratified by tacrolimus CV (>0.3 vs ≤0.3) and peak creatinine (>2.0 vs ≤2.0 mg/dL). High tacrolimus variability combined with elevated creatinine identified a cohort with 49.8% readmission rate compared to 24.8% in patients with low tacrolimus CV and preserved renal function (risk ratio 2.01, p<0.001, chi-square test) (Figure 4).

**Figure 4.**
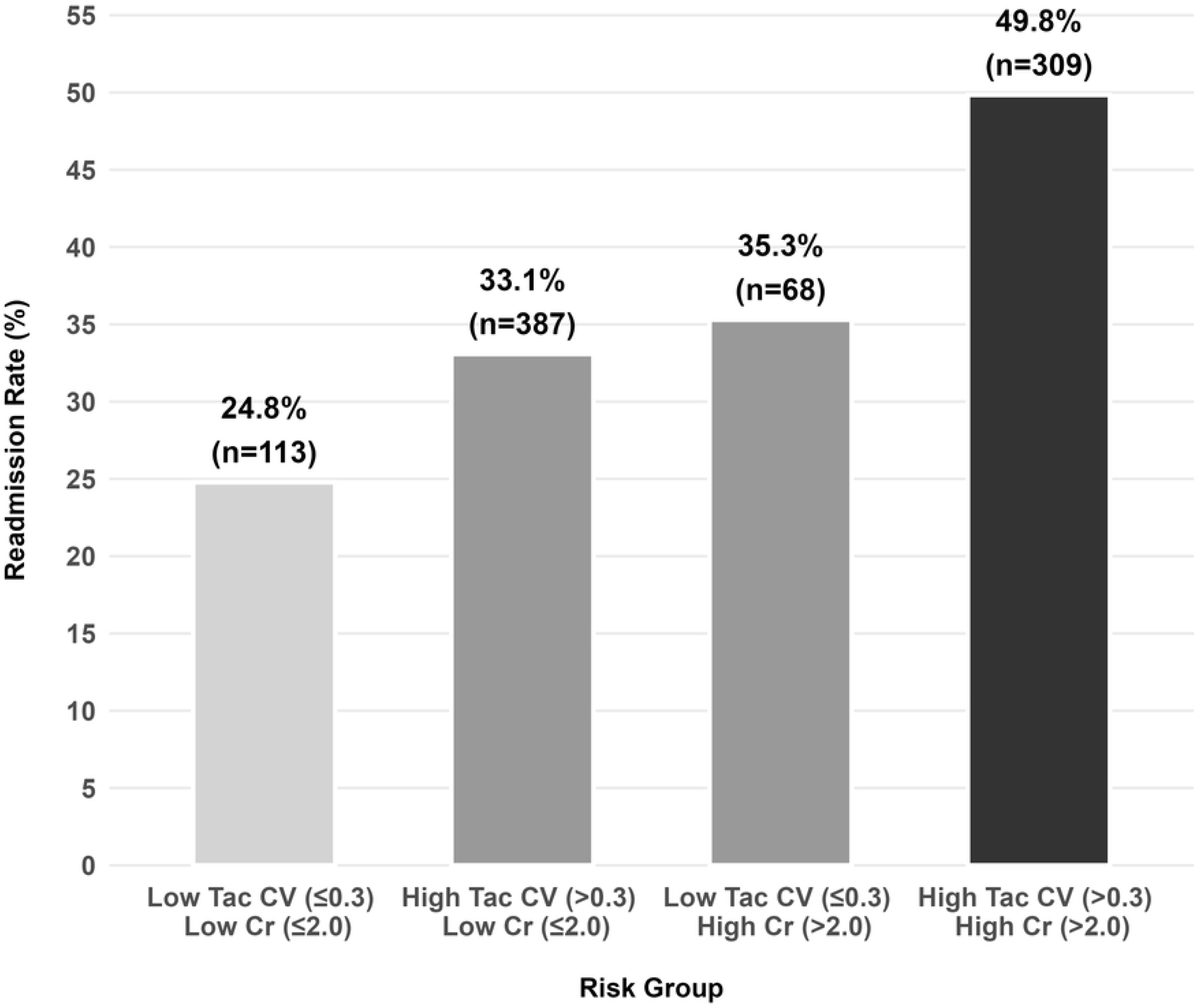
Stratified readmission rates. Readmission rates stratified by tacrolimus coefficient of variation (CV ≤0.3 vs >0.3) and peak creatinine (≤2.0 vs >2.0 mg/dL) during POD 0-7. Error bars represent 95% confidence intervals calculated using Wilson score method.

## DISCUSSION

Predicting unplanned hospitalization is difficult. This is true in both surgical and medical populations with systematic reviews demonstrating that even well-validated models rarely achieve C-statistics exceeding 0.70[13,14]. The conventional wisdom regarding the challenges in modeling readmission is that models are unable to capture problems arising de novo and not extensions of the hospitalization and fail to account for the environment within which post-operative recovery occurs.

Despite these challenges, unbiased models have a role in improving care. To that end, this work contributes to the understanding of post-liver transplant unplanned readmissions in three ways.

First, the study found that only 90-day readmission independently predicted one year mortality in liver transplant recipients. This contrasts with the post-kidney transplant readmission literature where readmissions within 30 days is the established mortality-predictive window[15]. The estimate provided is also the first in the liver transplant literature free of immortal time bias that complicates previous studies[16,17] which employed time-fixed analytical approaches that systematically underestimate the true association because patients must survive to be classified as readmitted, creating an immortal time that inflates survival in the admitted group.

The divergence from kidney transplantation likely reflects fundamental biological differences: whereas kidney allograft failure permits bridging with dialysis, there is no comparable rescue therapy for liver allograft dysfunction. The delayed mortality signal (Figure 1) suggests readmission-related complications manifest progressively rather than acutely, consistent with the protracted recovery trajectory of liver transplant recipients.

The second major finding is that accessible clinical data can be employed to create models with predictive accuracy similar to most post-surgical readmission models. In this study, the overall AUC for the POD 7 restricted model was 0.615 (95% CI 0.577-0.652) for a model incorporating only data available to POD 7. While the overall AUC reflects modest discrimination, this aligns with readmission prediction across surgical specialties[13] and exceeds the performance of many clinical risk scores in routine use. The POD 7 result is particularly interesting because there was a minimal difference in model performance between the two models even though it would be reasonable to consider that the inclusion of LOS and discharge location would be informative to a model. More importantly, the model’s value lies not in precise individual-level prediction but in identifying high-risk subgroups amenable to intervention which, in this study, were those with high tacrolimus variability and high peak creatinine who experienced a 2-fold readmission risk.

In both the complete data model and the POD 7 restricted model, the two highest ranked features were recipient BMI, and the interaction term between tacrolimus CV and peak serum creatinine. The association of recipient BMI with readmission contrasts with a prior study that reported obesity had no effect (or was protective against) readmission after liver transplantation[18]. The analysis, however, employed Cox regression models that did not account for immortal time bias with time-varying covariates. The effect of that bias is to inflate the survival estimate in the not-yet-readmitted group and explains the finding of a protective hazard ratio (HR 0.87).

Absent control for this bias, virtually any variable correlated with hospital length of stay (frailty, acute kidney injury, surgical complications, discharge disposition needs) will appear falsely protective in time-fixed models. Failure to address immortal time bias not only produces misleading hazard ratios but also misdirects clinical attention away from potentially modifiable risk factors.

The importance of the tacrolimus CV and peak creatinine interaction term to readmission is the third major finding of this study. The interaction is biologically plausible, aligns with the kidney transplant literature[19], and potentially actionable. This finding revealed a two-fold increase in risk for readmission in the highest strata of tacrolimus CV and peak serum creatinine. This finding invites interventions designed to reduce tacrolimus CV and avoid acute kidney injuries as a testable hypothesis to reduce readmission and, by extension, improve post-transplant survival when 70% of post-liver transplant patients are still hospitalized.

The single-center design offers both limitations and advantages. Multicenter data would be needed to confirm that the model’s discrimination generalizes across practice settings; however, the uniform laboratory platforms, immunosuppression protocols, and surgical techniques enhance measurement consistency, particularly critical for tacrolimus trough variability, where inter-institutional assay differences could confound multicenter analyses. This consistency may explain why the model identified biological signals (tacrolimus CV × peak creatinine) that aggregate datasets might obscure through measurement noise. More importantly, the single center approach aligns with the study objectives of identifying biological determinants of readmission risk rather than developing a deployable clinical risk calculator. Thus, this work should be evaluated as hypothesis-generating and providing a rationale for prospective intervention trials targeting tacrolimus variability.

An additional methodological strength is the manual validation of readmissions. Chart review of 50 randomly sampled readmissions confirmed all were genuinely unplanned (95% CI: ≥92.9% in full cohort), a validation rarely performed in transplant readmission research. This distinguishes post-liver transplant readmissions from other surgical populations where planned procedures often inflate readmission counts and obscure biological risk factors.

The models may also not apply to adult living donor liver transplants, as the cohort included only a very small number of domino transplants. The cohort also has a relatively small number of donors deceased by circulatory death (166, 18%). As the contribution of these donors reaches parity with donation after brain death, it will be necessary to retrain the model. Data relevant to organ recovery, for example, warm and cold ischemic time were not utilized as by choice because the significance of these measures is likely to change with the broader use of organ recovery and preservation technologies.

One of the most important opportunities for liver transplantation is to do for post-transplant predictions what MELD did for pre-transplant predictions. Within that context, this machine learning model leverages accessible clinical data while correcting pervasive methodological biases to identify modifiable determinants of readmission. The tacrolimus variability × acute kidney injury interaction represents a testable mechanistic hypothesis amenable to prospective validation through intervention trials, independent of whether the prediction model itself generalizes across centers. By establishing rigorous analytical methods, identifying actionable targets, and demonstrating feasibility of early risk stratification, this work provides both a methodological framework and biological foundation for designing interventions to reduce readmission and improve post-transplant outcomes.

## Data Availability

The dataset analyzed in this study contains protected health information extracted from the electronic health record. Due to the granularity of laboratory and clinical data combined with the small, single-center cohort, even de-identified data carry a meaningful risk of patient re-identification through triangulation with other available information. Public deposition is therefore not permitted under the approval granted by the Washington University School of Medicine Human Research Protection Office (HRPO). Requests for access to a restricted dataset should be directed to the Washington University HRPO (https://hrpo.wustl.edu/about-us/contact-us/), which will evaluate requests on a case-by-case basis in accordance with institutional data governance policy. The analysis code, covering feature engineering, model development, cross-validation, and statistical analysis, is publicly available at https://github.com/HepatologyLab/readmission_project without restriction. The code does not include data extraction or cleaning scripts, as these involve direct queries against the institution's electronic health record system and protected health information.

https://github.com/HepatologyLab

## ACKNOWLEDGEMENTS

The author acknowledges the assistance of Craig Cole and the Institute for Informatics, Data Science and Biostatistics (I^2^DB) for assistance in obtaining EHR data.

---

**Supplementary Figure 1.** Proportion of post-liver transplant patients hospitalized over time.

## REFERENCES

1. Axon RN, Williams MV. Hospital Readmission as an Accountability Measure. JAMA. 2011;305: 504. doi:10.1001/jama.2011.72

2. Li AH, Lam NN, Naylor KL, Garg AX, Knoll GA, Kim SJ. Early Hospital Readmissions After Transplantation: Burden, Causes, and Consequences. Transplantation. 2016;100: 713–718. doi:10.1097/TP.0000000000000917

3. Shankar N, Marotta P, Wall W, Albasheer M, Hernandez-Alejandro R, Chandok N. Defining readmission risk factors for liver transplantation recipients. Gastroenterol Hepatol N. 2011;7: 585–90.

4. Pereira AA, Bhattacharya R, Carithers R, Reyes J, Perkins J. Clinical factors predicting readmission after orthotopic liver transplantation. Liver Transpl. 2012;18: 1037–1045. doi:10.1002/lt.23475

5. Damazio B, Hao Q, Arenas JD, Riley TR, Hollenbeak CS. Risk factors for 30-day readmission following liver transplantation in Pennsylvania. J Liver Transplant. 2022;8: 100114. doi:10.1016/j.liver.2022.100114

6. Leslie Z, Leventhal T, Nguyen S, Leishman D, Espinoza C, Wise E, et al. Ninety-Day Readmission and Morbidity Following Liver Transplantation for MASLD. Clin Transplant. 2026;40: e70461. doi:10.1111/ctr.70461

7. Suissa S. Effectiveness of Inhaled Corticosteroids in Chronic Obstructive Pulmonary Disease: Immortal Time Bias in Observational Studies. Am J Respir Crit Care Med. 2003;168: 49–53. doi:10.1164/rccm.200210-1231OC

8. Lévesque LE, Hanley JA, Kezouh A, Suissa S. Problem of immortal time bias in cohort studies: example using statins for preventing progression of diabetes. BMJ. 2010;340: b5087. doi:10.1136/bmj.b5087

9. National Trends in Hospital and Physician Adoption of Electronic Health Records. In: ASTP Health IT Research & Analysis [Internet]. [cited 1 Mar 2026]. Available: https://healthit.gov/data/quickstats/national-trends-hospital-and-physician-adoption-electronic-health-records/

10. Amusa T, Okunola D, Izinyon O, Alabi A, Akinpeloye O. Strategies for Embedding Prediction Models in Clinical Decision-Making Workflows. Cureus. 18: e101185. doi:10.7759/cureus.101185

11. Chen T, He T, Benesty M, Khotilovich V, Tang Y, Cho H, et al. xgboost: Extreme Gradient Boosting. 2014. p. 3.2.1.1. doi:10.32614/CRAN.package.xgboost

12. R Core Team. R: A Language and Environment for Statistical Computing. R Foundation for Statistical Computing; 2025.

13. Kansagara D, Englander H, Salanitro A, Kagen D, Theobald C, Freeman M, et al. Risk Prediction Models for Hospital Readmission: A Systematic Review. JAMA. 2011;306: 1688. doi:10.1001/jama.2011.1515

14. Weinreich M, Nguyen OK, Wang D, Mayo H, Mortensen EM, Halm EA, et al. Predicting the Risk of Readmission in Pneumonia. A Systematic Review of Model Performance. Ann Am Thorac Soc. 2016;13: 1607–1614. doi:10.1513/AnnalsATS.201602-135SR

15. McAdams-DeMarco MA, Grams ME, King E, Desai NM, Segev DL. Sequelae of Early Hospital Readmission After Kidney Transplantation. Am J Transplant. 2014;14: 397–403. doi:10.1111/ajt.12563

16. Patel MS, Mohebali J, Shah JA, Markmann JF, Vagefi PA. Readmission following liver transplantation: an unwanted occurrence but an opportunity to act. HPB. 2016;18: 936–942. doi:10.1016/j.hpb.2016.08.003

17. Mumtaz K, Lee-Allen J, Porter K, Kelly S, Hanje J, Conteh LF, et al. Thirty-day readmission rates, trends and its impact on liver transplantation recipients: a national analysis. Sci Rep. 2020;10: 19254. doi:10.1038/s41598-020-76396-5

18. Ruck JM, Shui AM, Jefferis AA, Duarte Rojo A, Rahimi RS, Ganger DR, et al. Association of body mass index with post-liver transplant outcomes. Clin Transplant. 2024;38: e15205. doi:10.1111/ctr.15205

19. Borra LCP, Roodnat JI, Kal JA, Mathot RAA, Weimar W, van Gelder T. High within-patient variability in the clearance of tacrolimus is a risk factor for poor long-term outcome after kidney transplantation. Nephrol Dial Transplant Off Publ Eur Dial Transpl Assoc - Eur Ren Assoc. 2010;25: 2757–2763. doi:10.1093/ndt/gfq096

